# Resting-state causal brain connectivity in youth female athletes suggest sex-related differences following subacute concussion

**DOI:** 10.1101/2024.08.04.24311356

**Authors:** Julianne McLeod, Sahar Sattari, Dionissios T. Hristopulos, Karun Thanjavur, Naznin Virji-Babul

## Abstract

**Objective:** Youth male athletes show changes in resting-state causal brain connectivity following subacute concussion; however, little is known about how concussion alters causal brain connectivity in female youth. In this study, we compared resting-state causal brain connectivity in healthy and subconcussed females.

**Materials and methods:** Data from 11 concussed and 15 healthy control female athletes were included in this study. Five minutes of resting state eyes-closed EEG data were collected from all participants. SCAT5 data were also collected from all concussed participants. Causal connectivity was calculated from EEG source data. Network topology was evaluated using the degree assortativity coefficient, a summary statistic describing network structure of information flow between source locations.

**Results:** We observed three main results: 1) a qualitative difference in the spatial pattern of the most active connections, marked by posterior connectivity shifting in the concussed group, 2) an increase in the magnitude of connectivity in the concussed group, and 3) no significant difference in degree assortativity between the concussed and control groups.

**Conclusion:** Causal connectivity changes following concussion in females do not follow the same trends reported in males. These findings suggest a potential sex difference in injury response and may have implications for recovery.

## Introduction

Females are disproportionately affected by concussion in comparison to males, experiencing higher incidence rates, more severe cognitive dysfunction, higher post-concussion symptom scores, and longer recovery times (see 1,2 for reviews). Despite the lack of understanding behind these well-established sex-differences, females are underrepresented in concussion research. This issue was highlighted by D’Lauro et al. (3) in a review of consensus and position statements, revealing that male participants comprised 80% of the study populations referenced by the National Athletic Trainers’ Association, the International Conference on Concussion in Sport, and the American Medical Society for Sports Medicine. The most recent statement from the International Conference on Concussion in Sport (4) echoed this disparity, calling for more female-specific concussion research.

Functional brain connectivity changes following concussion have received increased attention in recent neuroimaging literature. It has been established that concussion disrupts functional connectivity with some suggesting that this disturbance to brain network communication plays a role in post-concussion symptoms like cognitive dysfunction (5–7). Connectivity changes are widely reported in the literature, with the default mode network (DMN) being a commonly investigated area. The DMN is a network of cerebral regions that show coordinated activity during wakeful rest and is associated with self-referential emotions and mind-wandering thoughts (8). Although not entirely undisputed, it is commonly reported that the acute phase after injury has increased functional connectivity (9–19). Reduced anterior connectivity paired with increased posterior connectivity has also been commonly reported (20–25). For detailed reviews, see (26,27).

Although there is substantial evidence that functional connectivity is disrupted by concussion, sex-specific findings are less commonly reported. Wang et al. (28) are one of the few groups to have compared functional connectivity between males and females with concussion. Their research indicates that males and females exhibit distinct patterns of functional connectivity following a mild traumatic brain injury. Specifically, males tend to show increased connectivity within motor, executive function, ventral stream, and cerebellum networks, but decreased connectivity within the visual network compared to females (28). The authors suggest that males and females may use different neural strategies for compensating and recovering from concussion-related disruptions, with males potentially being more vulnerable to certain types of neural dysfunctions. Interestingly, sex-differences in functional connectivity in those with persistent symptoms have also been reported (29). Both highlight the possibility of sex-specific casual connectivity changes post-concussion, warranting further investigation.

Increased functional connectivity, or hyperconnectivity, is a fundamental, purposeful response to mild, moderate, and severe traumatic brain injuries. Physical damage, such as axonal injury, is believed to trigger hyperconnectivity to restore network communication and allocate resources for structural repair (30–32). This phenomenon is more likely to occur in densely connected brain regions (31). While hyperconnectivity is adaptive in the short term, ultimately aiding in recovery, it can become harmful if sustained chronically. The increased metabolic cost associated with heightened neural communication can lead to resource depletion and potential neurodegeneration (30).

The pair-wise activity patterns extracted from functional connectivity analyses are non-directional, and therefore, do not provide insight into how neural information is transmitted between brain regions. A more powerful method that provides this directional information is known as causal/effective connectivity. Because neural signaling is inherently directional, examining causal connectivity changes post-concussion could provide more detailed insights into the underlying mechanisms of concussion.

A limited number of causal connectivity analyses have been employed in concussed populations (33–37). We previously investigated causal connectivity in a sample of adolescent male hockey players, finding significant differences between those with subacute concussion and healthy controls (33). Participants with subacute concussion showed symmetrical bilateral patterns with reduced central-parietal activity and additional frontal-temporal connections. Another study found increased posterior-to-anterior effective connectivity following concussion in sample of male high school football players (34). In a pediatric case-control sample, post-concussion effective connectivity within the DMN showed increased inter- and intra-hemispheric anterior connectivity, with unique connections from the orbitofrontal cortex to parietal regions (35). Acute concussion in adults has been associated with decreased connectivity from the left middle frontal gyrus to various areas in the temporal, frontal, and insular regions, from the left insula to areas within the frontal and central regions, and from the right insula to the left superior frontal gyrus. Increased connectivity was observed from the left anterior cingulate cortex to areas of the frontal lobe and the left insula, as well as from the right insula to the left superior temporal gyrus (36,37). Hristopulos et al. (33) and Reddy et al. (34) included only male participants. Vaughn et al. (35) and Li et al. (36,37) included both males and females, but did so in pediatric and adult populations. There is a significant gap in the literature regarding post-concussion causal connectivity in female youths. To address this gap, we examined resting-state causal connectivity in female athletes aged 15 to 24 with subacute concussion (≤1 month) and no reported history of concussion. We applied Information Flow Rate to source-reconstructed EEG data to obtain causal connectivity network maps for both concussed and control groups, aiming to investigate the network changes following concussion. To our knowledge, this is the first study to explore causal connectivity in a concussed-control design within an entirely female athlete youth sample.

The secondary objective of this study was to explore whether subacute concussion is associated with differences in resting-state network topology, as measured by the Degree Assortativity Coefficient (r_w_). Our previous research found youth athletes with subacute concussion to have significantly higher global assortativity compared to healthy controls (33). Increases in local assortativity have also been reported in adults with acute concussion, particularly in the frontal regions (26). Churchill et al. (38) report different results, finding that university athletes within 1 week of concussion to have lower r_w_ values compared to healthy controls. Repeated measures showed that r_w_ linearly increased across recovery, raising the possibility that tracking this metric could offer an objective way to evaluate injury recovery, a tool that is currently lacking. Apart from Churchill et al. (38), these studies included predominantly or entirely male samples. In this study, we address this sex and age gap by investigating global changes in r_w_ following concussion in female youth athletes. This information may be particularly relevant for contributing to information about sex-specific diagnostic and management strategies.

## Materials and methods

### Participants

Twenty-six right-handed, female athletes between 15 and 24 years of age were recruited for this study. Participants were recruited from various contact and non-contact sports teams, including soccer, rugby, and swimming. Healthy control participants reported no history of concussion. Concussed participants were within 30 days of injury, reported a diagnosis of concussion from a physician or team doctor, were symptomatic at time of study enrolment, and reported no medical clearance to return to play (RTP). Exclusion criteria for all participants included focal neurologic deficits and/or diagnosis with a neurological condition. This study was approved by the University of British Columbia Clinical Research Board (H17-02973) in accordance with the Helsinki declarations. All participants provided written informed consent before participating.

### Concussion symptom assessment

The Sport Concussion Assessment Tool 5 (SCAT5) was used for assessing the number and severity of concussion symptoms. The SCAT5 is a standardized sport concussion assessment tool that is used to evaluate the presence and severity of concussion symptoms (39). Respondents are asked to rate the severity of a series of 22 symptoms on a 6-point scale ranging from 0 (none) to 6 (severe). Two scores are generated from the questionnaire: total number of symptoms and total severity score. The SCAT5 data were collected at the time of EEG recordings.

### EEG acquisition

Five minutes of resting-state eyes-closed EEG data were collected for all participants using a 64-channel HydroCel Geodesic Sensor Net (EGI, Eugene, OR) connected to a Net Amps 400 high-impedance amplifier. Figure 1 shows the procedure of EEG preprocessing and analysis. The vertex (Cz) served as the reference point during the recording setup. Data were recorded at a 500 Hz sampling rate, with scalp electrode impedances typically less than 50kΩ.

**Figure 1.**
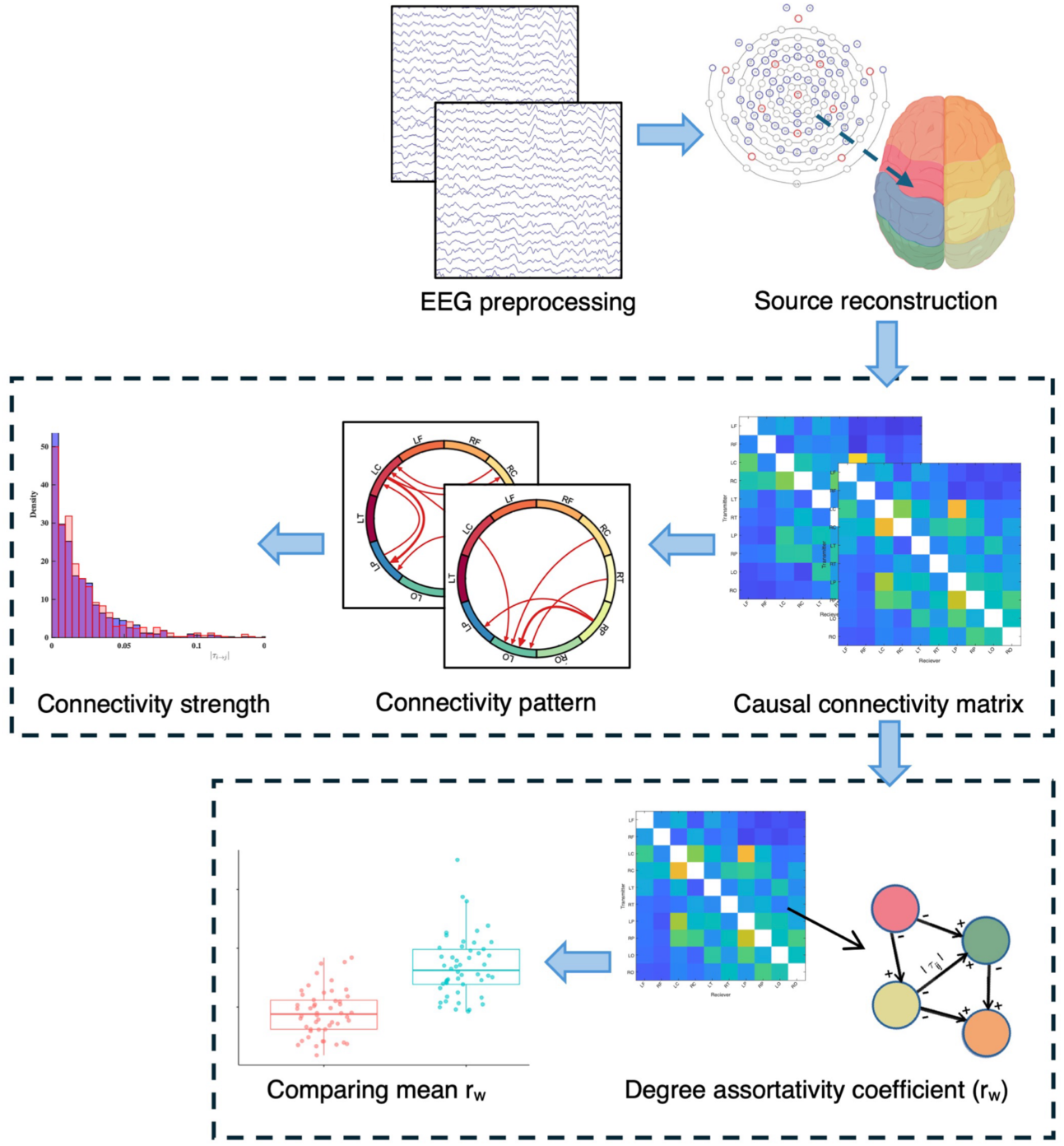
EEG preprocessing and analysis procedure.

### EEG preprocessing and source reconstruction

Raw EEG data were preprocessed using EEGLAB in MATLAB (40). Data were re-referenced to the average of all channels, downsampled to 250 Hz, notch filtered, and bandpass filtered between 0.5 and 50 Hz. Non-brain artifacts (i.e., noise caused by participant motion) identified through Independent Component Analysis (ICA) and visual inspection were removed.

Clean EEG sensor-level data were transformed to source space using Brainstorm in MatLab (41). An MRI head template (ICBM152) was used for head modelling (42). The head model was divided into three sections (scalp, skull, brain) for forward modelling. The source space was constrained to the cortex. Minimum Norm Estimate (MNE) and sLORETA were used for inverse modelling (43,44). The cortical surface was symmetrically divided into ten regions of interest (ROIs) using the Desikan-Killiany atlas (45). The ten predetermined ROIs were as follows: left frontal (LF), right frontal (RF), left central (LC), right central (RC), left parietal (LP), right parietal (RP), left temporal (LT), right temporal (RT), left occipital (LO), and right occipital (RO).

Each of the ten source regions generated a distinct time series, assumed to reflect the sum of electrical activity occurring in that specific brain area. The resulting clean source reconstructed EEG data were used to calculate causal connectivity.

### Causal connectivity

Causal connectivity was quantified using Liang-Kleeman’s Information Flow Rate (IFR) (46). IFR provides a framework for measuring the transfer of information in dynamic systems and is particularly suitable for time series data (47). The theorem is briefly described here, and a comprehensive review of the framework is reported in (46).

The covariance between two source regions, *v*_*i*_ (transmitting information) and *v*_*j*_ (receiving information), is computed with the *sample cross-covariance* formula. This calculation quantifies how much time series *v*_*i*_ and *v*_*j*_ are correlated over time. Specifically, it assesses how fluctuations in *v*_*i*_ are associated with the fluctuations in *v*_*j*_, examining deviations from their average values over time. For dipoles located at *v*_*i*_ and *v*_*j*_, the sample cross-covariance coefficient is denoted as *Ĉ*_*i,j*_. The magnitude (strength of electrical activity) at source location *v*_*i*_ and at time point *n* is *v*_*i*,*n*_; for source location *v*_*j*_, *v*_*j,n*_. The computation of the sample cross-covariance uses the following formula:

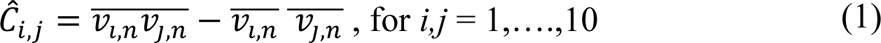

where 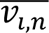 and 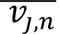 represent the mean values of *v*_*i*,*n*_ and *v*_j,*n*_, respectively.

The Pearson correlation between time series *v*_*i*_ and *v*_*j*_ (*r̂*_*i,j*_) is a normalized measure of the linear relationship between the activities of two brain regions. Both *Ĉ*_*i,j*_ and *r̂*_*i,j*_ are non-directional and *r̂*_*i,j*_ is commonly reported as a measure of functional connectivity. The *cross-correlation coefficient* between *v*_*i*_ and *v̇*_*y*_ (the temporal derivative of *v*_*j*_) over time and is denoted as *r̂*_*i,dj*_ This quantifies how the activity in one brain region relates to the rate of change in the activity of another region. The formulas to estimate *r̂*_*i,j*_ and *r̂*_*i,dj*_ are:

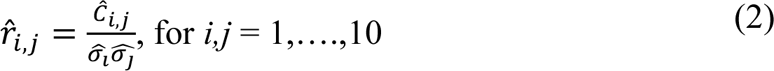

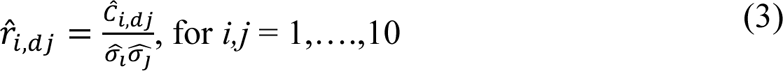

where 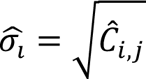 is the sample standard deviation of *v*_*i*_ and *Ĉ*_*i*,*dj*_ is the sample covariance of *v*_*i*_ and first derivative of *v*_*j*_.

The information flow rate from the time series *v*_*i*_ to *v*_*j*_(*T*_*i*→j_) is calculated using the *Liang-Kleeman coefficient* formula. To evaluate the significance of the information flow, the normalized information flow rate (*τ_i→j_*) is calculated. The coefficient *τ_i→j_* measures the proportion of total entropy rate change in *v*_*j*_attributed to *v*_*i*_. Computation of *τ_i→j_* uses a normalization factor (*Z*_*i*→j_) that is further described in Liang (48). Higher |*τ_i→j_*| values signify greater information transfer between regions. A threshold of 0.05 has been used to identify *active* connections, indicative of meaningful influence between brain regions.

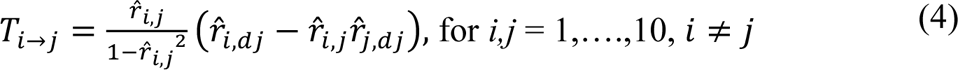

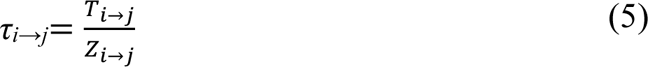

The resulting output from the casual connectivity computation for a single participant is a binary weighted directed network matrix with the magnitude (strength) of each connection corresponding to |*τ_i→j_*|. Connectivity matrices for individuals were averaged to produce a single matrix for concussed and control groups, followed by a between-group comparison.

### Degree assortativity

As a secondary outcome measure of brain connectivity, we compared the degree of assortativity between the concussed and control groups. Assortativity is a summary statistic of a network’s topological structure. It was originally conceptualized by Newman et al. (49) and later studied in the context of brain connectivity. It is a graph theory metric that calculates the tendency of nodes in a network to connect with other nodes of similar degree. Assortativity is expressed on a scale ranging from −1 (perfectly disassortative) to 1 (perfectly assortative) (50).

We calculated the degree assortativity coefficient (r_w_) using an adaptation of Newman’s original formula for weighted, directed networks, as described by Rubinov and Sporns (51). The r_w_ is the Pearson correlation between the degree of nodes at either end of an edge (50). The 10 predefined ROI source regions are the nodes, and the |*τ_i→j_*| are the weighted edges. The binary matrix generated from the casual connectivity calculation is used to determine the weight of each node. Each node is assigned an in-degree and an out-degree. The in-degree is calculated from the sum of the |*τ_i→j_*| for all incoming connections. The out-degree is calculated from the sum of the |*τ_i→j_*| for all outgoing connections. We use the pearson.W.m function from the Octave networks toolbox in MatLab to calculate r_w_ (52).

The r_w_ was calculated for each subject based on their individual matrix. Mean r_w_ was calculated by averaging all individual r_w_’s. Between-group comparison of mean r_w_ was conducted using an independent samples t-test. The independent samples effect size using Cohen’s d was reported.

### Statistical analyses for casual connectivity

Descriptive statistics were computed for the demographic data and SCAT5 scores. All tests were conducted at 5% significance. The analyses were completed using SPSS software (version 29.0.20.0).

The statistical significance of all mean connections was assessed. Using non-parametric permutation testing, we tested the null hypothesis that there was no information flow between *v*_*i*_ and *v*_*j*_. For each participant, the transmitter time series was permuted 100 times to generate 100 permuted |*τ_i→j_*| values for each connection. The significance threshold was set by calculating the 5th and 95th percentiles of these permuted distributions derived from the permutations.

The magnitudes of |*τ_i→j_*| were compared between both groups. One distribution per group was generated using |*τ_i→j_*| values for each of the 90 connection pairs between the 10 ROIs per individual in each group. Hence, meaning that the distributions consisted of 15 × 90 = 1350 |*τ_i→j_*| values in the control group and 11 × 90 = 1080 |*τ_i→j_*| values in the concussed group. Differences in the shape and central tendencies between the two distributions were assessed using the Kolmogorov–Smirnov and Kruskal– Wallis tests. 95% confidence intervals were reported for the coefficient of variation (COV), skewness, and kurtosis corresponding to further describe differences between the distributions. One hundred thousand (100,000) subsamples of six individuals were generated from random subsampling of each group, and plots were generated to visualize the resulting COV, skewness, and kurtosis of each distribution.

## Results

### Clinical and demographic data

Table 1 presents the descriptive statistics for the concussed and control groups. Age for the entire sample (N=26) ranged from 15 to 24 years: *n*=11 with subacute concussion (M = 20; SD = 2.46) and *n*=15 healthy controls (M = 21 years; SD = 1.88). All concussed participants reported a diagnosis of concussion and averaged 10 days post-injury (SD = 6.85). On average, the concussed group reported 14.88 (SD = 5.48) symptoms with a severity of 34.88 (SD = 22.53; range = 7-71) on the SCAT5.

**Table 1.** Demographic and concussion symptom assessment data.

| | Control ($n=15$) | Concussed ($n=11$) |
| --- | --- | --- |
| Age (years) | $21 \pm 1.88$ | $20 \pm 2.46$ |
| Time since injury (days) | - | $10 \pm 6.85$ |
| Diagnosed concussions to date | - | $2.25 \pm 1.39$ |
| SCAT5, total number of symptoms | - | $14.88 \pm 5.48$ |
| SCAT5, severity of symptoms | - | $34.88 \pm 22.53$ |

*All values are presented as mean ± standard deviation.*

### Between-group differences in the causal connectivity matrices

#### Pattern of information flow

The spatial distributions of the ten strongest connections for the control and concussed groups are compared in Figure 2. The rank ordered magnitudes and directions of these connections are listed in Table 2. For simplicity, only the ten strongest connections were qualitatively assessed, and show clear differences between the concussed and control groups.

**Figure 2.**
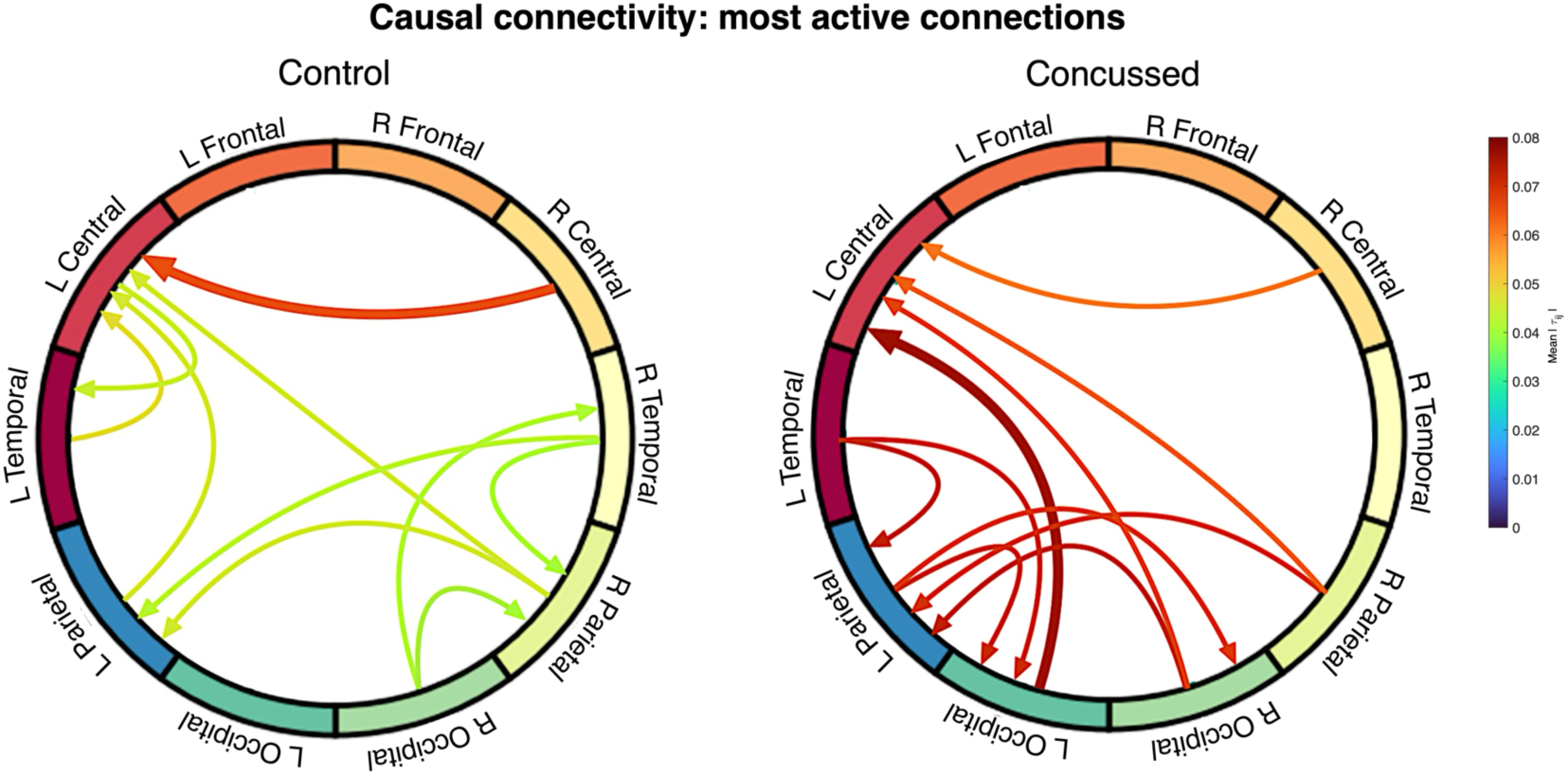
Spatial distribution of the strongest connections. The graph layout reflects the superior view of the cortical surface. Arrows represent the direction of causality, with arrow color corresponding to the magnitude (|*τ_i→j_*|) of the connection.

**Table 2.**
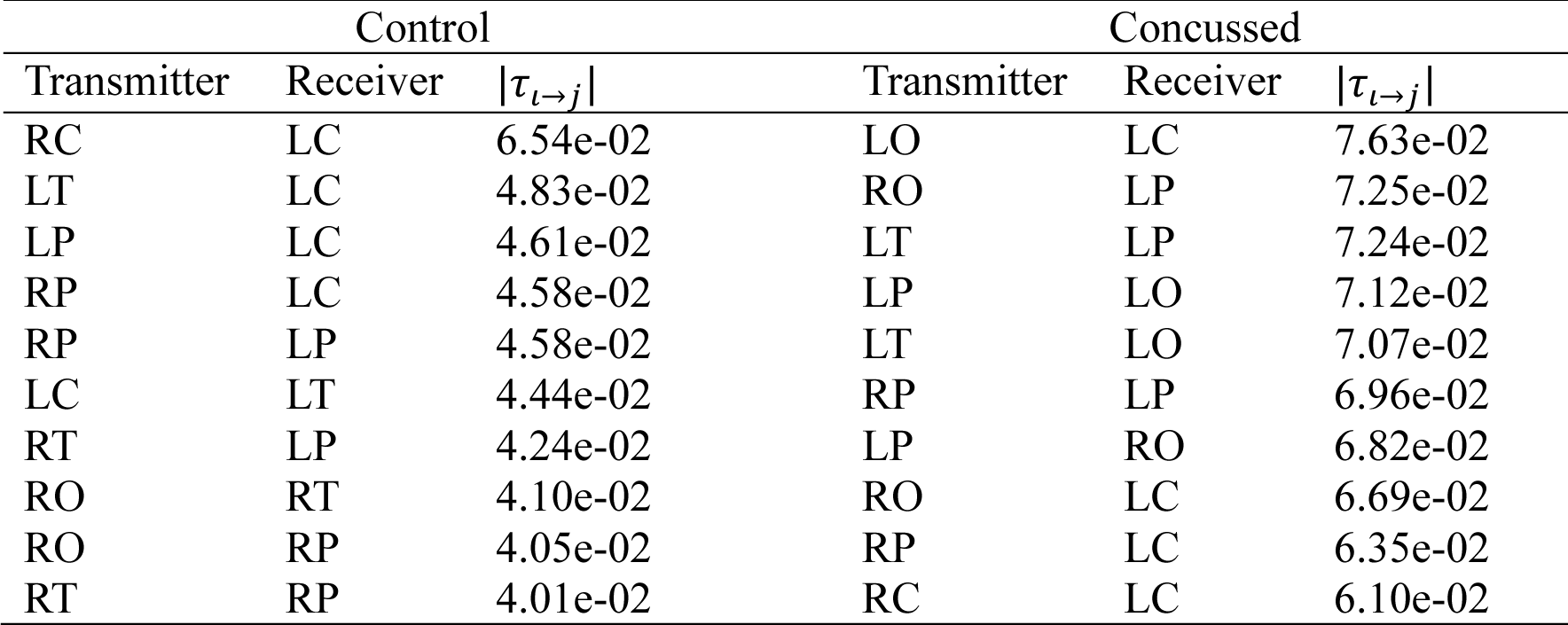
Top ten strongest connections ranked by the magnitude of mean|*τ_i→j_*| values for control and concussed groups.

| Control |  |  | Concussed |  |  |
| --- | --- | --- | --- | --- | --- |
| Transmitter | Receiver | $\vert \tau_{i \rightarrow j}\vert $ | Transmitter | Receiver | $\vert \tau_{i \rightarrow j}\vert $ |
| RC | LC | 6.54e-02 | LO | LC | 7.63e-02 |
| LT | LC | 4.83e-02 | RO | LP | 7.25e-02 |
| LP | LC | 4.61e-02 | LT | LP | 7.24e-02 |
| RP | LC | 4.58e-02 | LP | LO | 7.12e-02 |
| RP | LP | 4.58e-02 | LT | LO | 7.07e-02 |
| LC | LT | 4.44e-02 | RP | LP | 6.96e-02 |
| RT | LP | 4.24e-02 | LP | RO | 6.82e-02 |
| RO | RT | 4.10e-02 | RO | LC | 6.69e-02 |
| RO | RP | 4.05e-02 | RP | LC | 6.35e-02 |
| RT | RP | 4.01e-02 | RC | LC | 6.10e-02 |

In the control group, the top ten connections are widespread. The strongest connections are predominantly within anterior regions, with the top connection transmitted from the right to left central area. The left central region is a primary hub for receiving information. Three inter-hemispheric connections are observed, including right-to-left communication between temporal and parietal areas. Left sided intra-hemispheric connections are more focused in anterior regions (LC➔LT, LT➔LC, LP➔LC), while right sided intra-hemispheric connections are more localized the posterior region (RO➔RT, RO➔RP, RT➔RP).

In the concussed group, connectivity shifts to more posterior and left-lateralized regions. The strongest connection is between the left occipital to the left central region. The left occipital and parietal area are primary hubs for incoming and outgoing connections. There is an increase in strong inter-hemispheric connections (RO➔LP, RO➔LC, RP➔LP, RP➔LC, RC➔LC) and a decrease in right intra-hemispheric information flow.

Common connections for both groups include RC➔LC, RP➔LC, RP➔LP. Unique to the control group are connections transmitted from RT. Information transmitted from the right occipital is consistent between the two groups, but its targets differ: RO➔RT and RO➔RP in the control group RO➔LC and RO➔LP in the concussed group. The concussed group shows more information flow to and from the left occipital (LO➔LC, LT➔LO, LP➔LO) and left parietal (incoming connections from the LT, RO, and RP and outgoing to RO and LO). The left temporal transmits information posteriorly in the concussed group (LT➔LO, LT➔LP) compared to anteriorly in the control group (LT➔LC). The left central region is a main receiver of inter- and intra-hemispheric information in both groups, but only the control shows a strong outgoing connection.

#### Magnitude of information flow rates

Figure 3 presents binary, weighted matrices showing the mean absolute normalized information flow rates (|*τ_i→j_*|) for the concussed and control groups. Overall, connection strengths (|*τ_i→j_*|) were higher in the concussed group. The difference in magnitude of |*τ_i→j_*| is visible in the plots due to the color bar scale. The concussed group matrix shows higher magnitudes (red), which is particularly clear in Figure 3B, where only the ten strongest connections are shown.

**Figure 3.**
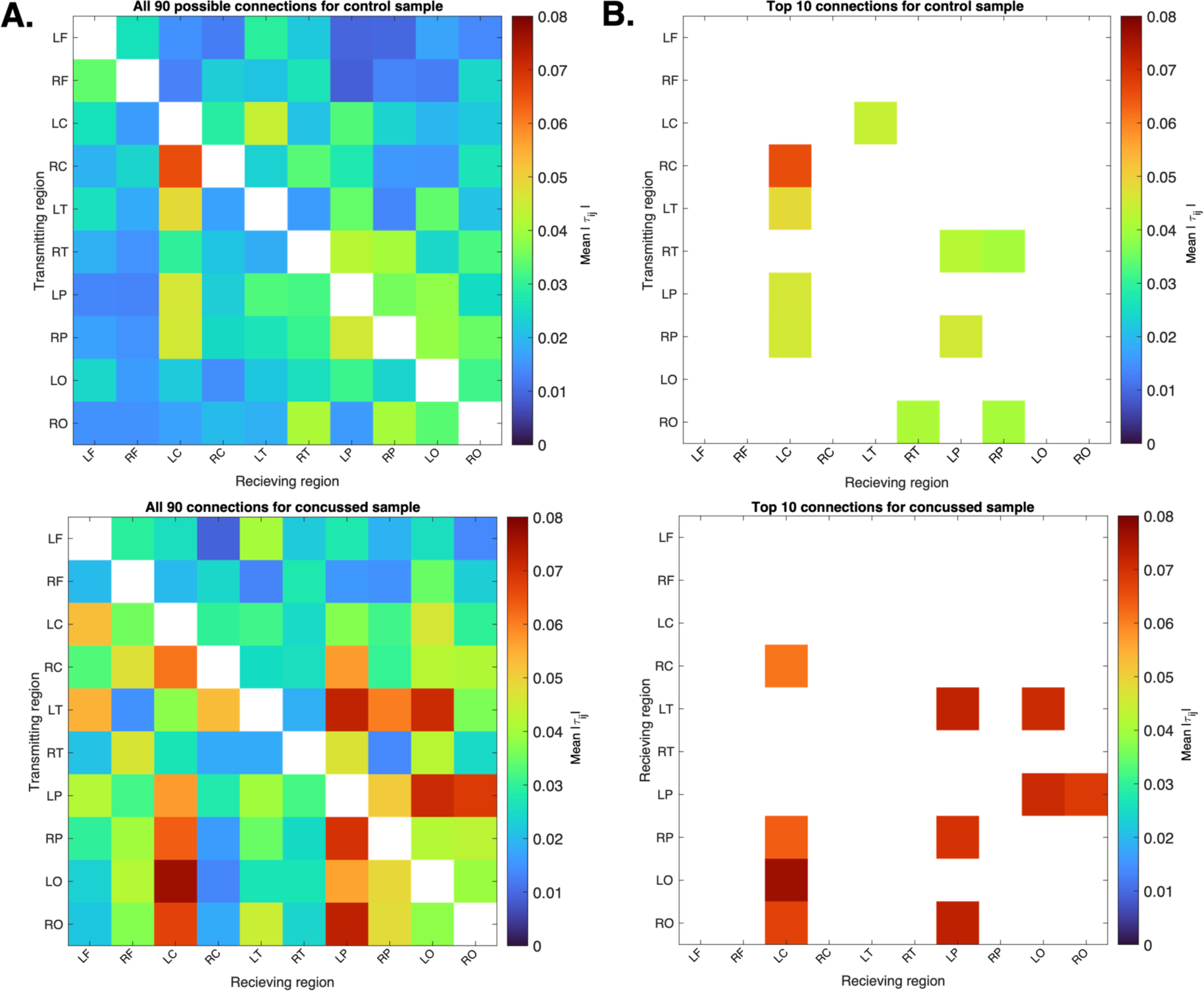
Causal connectivity matrices for the control (top) and concussed (bottom) groups. The maps illustrate the mean |*τ_i→j_*| for (**A**) each of the 90 possible pairs of the ten source ROIs, and (**B**) the ten strongest connections for each group.

The ten strongest connections are listed in Table 2 and illustrated in Figure 3B. For the list of all 90 rank ordered connections refer to Appendix A. The magnitudes of the top ten connections in the control group ranged from 4.01e-2 to 6.54e-2, with only the top connection (RC➔LC) exceeding the *active* threshold of 0.05. The concussed group’s top ten connections ranged from 6.10e-2 to 7.63e-2, with 19 connections exceeding the *active* threshold.

### Between-group differences in the degree assortativity coefficient

The mean r_w_ was higher in the concussed group (*M* = 0.41, *SD* = 0.26) compared to the control group (*M* = 0.27, *SD* = 0.22) (Figure 4). However, an independent samples t-test assuming unequal variances found no statistically significant difference between the groups, *t*(19.79) = 1.39, *p* = .18. The mean difference was 0.13 (SE = 0.09), with 95% CI equal to [−0.07, 0.33], with Cohen’s *d* indicating a small effect size (i.e., the difference in the means is small compared to the variability), *d* = 0.24, with 95% CI [−0.24-1.35].

**Figure 4.**
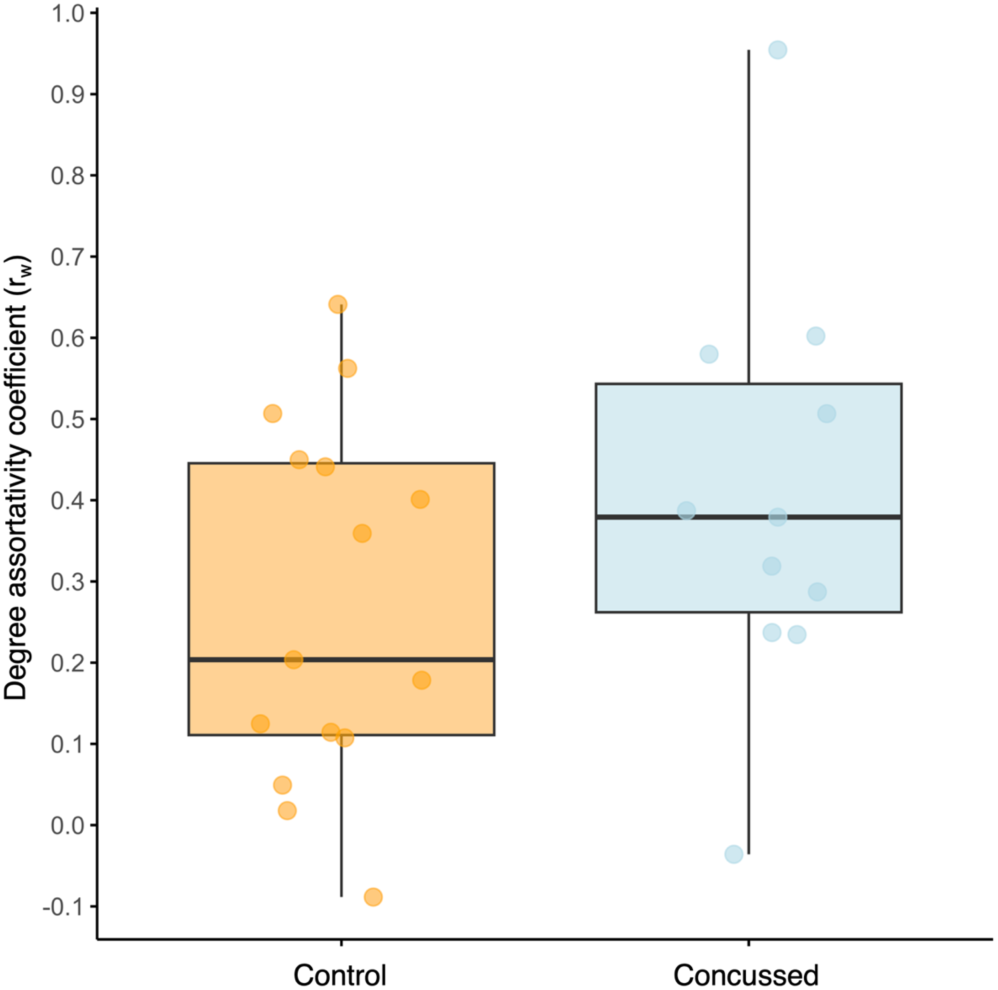
Boxplot comparing the degree assortativity coefficient (r_w_) between the control and concussed groups.

### Statistical analyses for causal connectivity

First, we determine the statistical significance of all 90 mean connections in each group. The 95th percentile of the |*τ_i→j_*| randomized distribution derived by means of permutation testing were 1.38e-05 for the control group and 1.75e-05 for the concussed group. Since all mean connections exceeded these thresholds, statistical significance was confirmed for both groups.

Next, we generate a probability density histogram consisting of two distributions comprising all individuals in each group. The probability density histogram (Figure 5A) shows that the control group was more likely to have lower mean |*τ_i→j_*| values compared to the concussed group. The Kolmogorov-Smirnov (K-S) and Kruskal-Wallis (K-W) test indicate that the distributions are significantly different (*p* = 1.61e-05 and *p* = 1.50e-03, respectively). Table 3 lists the 95% confidence intervals for coefficient of variation (COV), skewness, and kurtosis for the distributions.

**Figure 5.**
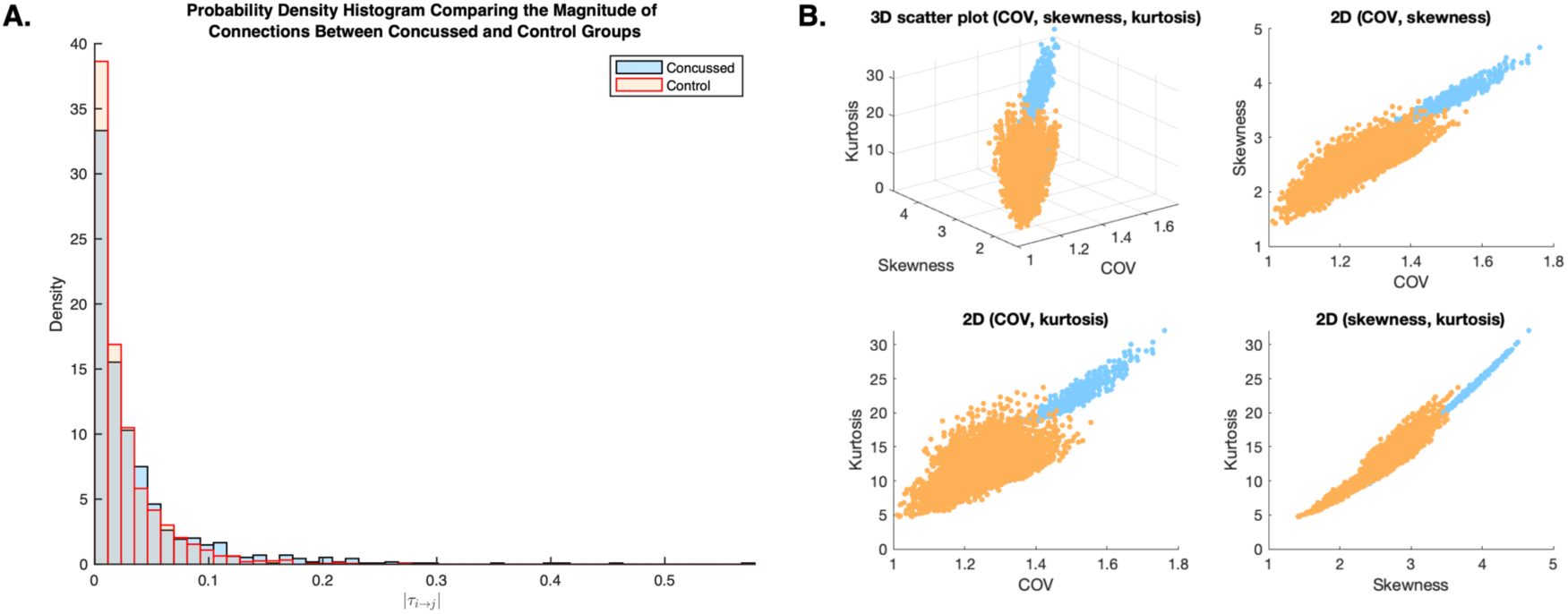
Comparing |*τ_i→j_*| distributions between the concussed and control groups. (**A**) Probability density histogram comparing two distributions (control and concussed). All individuals are included in each group. The density (y) of individual connections at each given magnitude (x) illustrates the overall strength of causal connections in each group (**B**) Scatter plots of COV, skewness and kurtosis derived from 100,000 subsamples, each comprising 6 randomly selected individuals. The COV, skewness and kurtosis are based on all the connections and individuals in each subsample.

**Table 3.** Statistics corresponding to the probability density histogram distributions of |*τ_i→j_*| connections for the control and concussed groups.

| Group | COV | Skewness | Kurtosis |
| --- | --- | --- | --- |
| Concussed | [1.13-1.32] | [1.97-2.81] | [7.05-13.79] |
| Control | [1.03-1.28] | [1.48-2.37] | [5.20-10.38] |

*Values are presented as 95% confidence intervals.*

Lastly, we select a subgroup of individuals from each group and derive subsampling distributions of information flow rates to further evaluate characteristics of each group’s distribution (Figure 5B). Even though the distributions derived from subsampling follow the same orientation, the distributions are fairly well separated and the shapes generally differ between the two groups. Although the two groups are not completely separated in this three-dimensional feature space, the observed behavior is in overall agreement with the results from the K-S and K-W tests, which indicated that the distributions are distinct from one another.

## Discussion

In this study, we investigated resting-state causal connectivity and network topology in female youth athletes with subacute concussion and healthy controls. We applied Information Flow Rate to source-reconstructed EEG data in order to obtain binary causal connectivity matrices and to compare connectivity patterns between the groups. As a secondary outcome measure, we calculated the degree assortativity coefficient (r_w_) to assess the network topology of each group. Our aim was to evaluate changes in causal connectivity in a small cohort of concussed female youth athletes compared to an age- and sex-matched non-concussed group. Below, we explain the significance of each observation and compare with relevant literature on males.

Our findings revealed distinct differences in resting-state brain causal connections between concussed and control female youth athletes. Specifically, we found three key results: (1) the spatial distribution of the strongest connections was different between the two groups, (2) the concussed group had significantly higher magnitudes of information flow, and (3) network assortativity was not significantly different.

### 1) Qualitative shift in the pattern of information flow

The control group had widespread connectivity with relatively equal inter- and intra-hemispheric connections, while the concussed group’s strongest connections were mainly localized within posterior areas and were left-lateralized. Our findings partially align with Vaughn et al. (35). Both studies report more widespread activity in healthy controls, but while Vaughn et al. (35) observed greater anterior activity in the concussed group, we found more posterior activity.

Vaughn et al. (35) also observed increased causal connectivity from the left orbitofrontal cortex to the right parietal regions following concussion. Similarly, we observed an increase between the right parietal and left central region in our concussed sample, though in the opposite direction (RP→LC). The opposite directionality may be attributed to age differences, as Vaughn et al. (35) studied a younger sample. Michels et al. (53) support this age-related difference, noting anterior-to-posterior information flow in children and posterior-to-anterior in adults. The reason for increased connectivity between these regions post-concussion is still debated. Vaughn et al. (35) found that stronger orbitofrontal-to-parietal connectivity correlated with fewer post-concussion symptoms. In adults, increased frontal connectivity has similarly been linked to fewer symptoms (17). Our study shows that a similar pattern of increased frontal-parietal connectivity exists in youth females following concussion. However, further research is needed to explore its relationship with post-concussion symptoms.

We observed reductions in certain connections in concussed females, which might align with the reductions reported by Li et al. (36,37). Specifically, connections such as LC→LT, RC→LC, and LC→RT were stronger in the control group compared to the concussed group. Li et al. (36,37) reported reduced connectivity from the left prefrontal cortex to the left middle temporal gyrus, from the left insula to regions of the prefrontal cortex, and from the left insula to the right rolandic operculum. Reductions in these areas could be associated with symptoms such as reduced information processing, poor working memory, emotional dysregulation, and poor cognitive control. While these findings suggest similar trends, methodological differences like Li et al.’s use of fMRI with seed-based ROIs compared to our EEG study with region-based cortical ROIs, thus making it challenging to draw strong conclusions. Future research should validate these findings and explore causal connectivity changes during recovery in youth females.

### 2) Increased magnitude of information flow

Greater magnitudes of information flow (stronger connectivity) in the concussed group compared to the control group align with numerous reports of increased functional connectivity in the acute phase of concussion (9–19). The significantly higher magnitudes of information flow are suggestive of hyperconnectivity, indicating that the concussed female youth brain exerts more effort at rest compared to a healthy, uninjured brain.

Hyperconnectivity in the acute phase of injury might be an adaptive response, potentially occurring to re-establish network communication or to allocate resources for repair (30–32) of microstructural lesions and/or neurometabolic alterations (54–56). While potentially beneficial in the short term, chronic hyperconnectivity has a high metabolic cost that can lead to resource depletion and neurodegeneration (30). Disruptions in functional connectivity have been observed in adult and pediatric populations with persistent symptoms (6,29,57–60), as well as recovered athletes that have been cleared for RTP (22,61–64). Data in female youth is lacking, necessitating further studies to validate our observations and to explore how increased information flow shifts along recovery.

### 3) No difference in network assortativity

The concussed group had a slightly higher mean degree assortativity coefficient (r_w_) compared to the control group, however this difference was not statistically significant. This suggests that the two groups did not differ in topological structure and since the mean r_w_ exceeded 0 in both groups, the networks were considered assortative.

The degree assortativity coefficient is reflective of network robustness and interconnectivity (65). Highly assortative networks are less likely to become disconnected after trauma, whereas disassortative networks are more vulnerable (50). Greater assortativity suggests resilience and might be an adaptive response to injury (51). Our concussed cohort did not show this adaptive response, which could be an indication that the female response to concussion may differ from what has been reported in the literature (which is primarily on males).

Although Churchill et al. (38) reported decreased assortativity acutely after concussion, their study included an equal distribution of university-aged males and females but did not report female-specific assortativity scores, possibly obscuring the results. To our knowledge, there are no other studies that have evaluated degree assortativity coefficient following concussion in females, underscoring the need for more research in this area.

#### Possible sex differences

Behaviorally, females have been reported to take longer to recover, and experience a greater symptom burden in comparison to males (1,2,66–68); however, recent work by Bennett et al. (69) and others (70,71) is starting to reveal that males and females have quite different responses to concussion. Bennett et al. (69) investigated changes following sub-concussive hits in male and female boxers and found that the linear relationship between brain volume loss and sub-concussive hits in males was significantly more severe compared to females. Others (70,71) have found that males show greater white matter disruption following concussion compared to female athletes. These findings hint at the possibility that concussion in males leads to more pronounced underlying neuropathological changes compared to females. Below we discuss our findings in females in relation to what has been reported in males.

In our previous work in adolescent males, we observed post-concussion causal connectivity to be primarily antero-centric with fewer posterior connections (33). This is seemingly opposite to what we observed in the present study, where the strongest connections in the female concussed group were primarily posterior. Sex differences in functional connectivity studies have been reported, with Wang et al. (28) reporting differences acutely after concussion and Shafi et al. (29) reporting more chronic differences in an investigation of males and females with post-concussion syndrome. To date, no study has evaluated sex-specific differences in causal connectivity following concussion. Our findings in females, contrasted with our previous work in males, suggest the likelihood of such differences and underscore the importance of further investigation in age-matched males and females.

The change in magnitude |*τ_i→j_*| that we observed in females showed an opposing trend to that of concussed-control males reported in Hristopulos et al. (33). Concussed females exhibited increased magnitudes of information flow, while concussed males had decreased magnitudes. This trend is in line with Churchill et al. (72) who reported decrease of cerebral blood flow to decrease following concussion in males in contrast with an increase in females. Additionally, the between-group magnitude difference was less distinct in females as compared to the males, as evidenced by overlapping confidence intervals and slightly overlapping COV, skewness, and kurtosis distributions. This could suggest that differences in females are less distinct than those observed in males, aligning with reports by Bennett et al. (69) and others (70,71).

Unlike Moreira da Silva et al. (73) and Hristopulos et al. (33), who reported significant increases in assortativity following acute concussion, we did not observe a significant change in assortativity in our concussed female sample. Importantly, only 16% of the concussed sample in Moreira da Silva et al. (73) was female, while Hristopulos et al. (33) only included males. Our opposing findings might suggest a sex-specific response to injury.

The biological causes of the observed differences between males and females in terms of causal connectivity changes post-concussion are unclear. Hormonal differences are often acknowledged in discussions regarding sex-differences in concussion, yet we still have a poor understanding of how hormones might influence outcomes and even less of an understanding in the context of brain connectivity. Recently, we found cycle phase to be associated with significant changes in information flow patterns and degree assortativity in healthy females (74), highlighting the importance of considering brain-hormone relationships. Sex differences in neuropathology, like females having reduced total brain volume, regional volume differences, smaller axon diameter, and reduced thickness of the corpus callosum (75,76), may play a role in how causal connectivity changes after injury. However, further longitudinal, multimodal studies are needed to better understand these sex-specific causal connectivity changes and their implications for concussion outcomes.

#### Limitations and future directions

This study is not without limitations. First, we have a small sample size. Larger studies are needed to validate our findings. We only collected data in the subacute phase of injury, and therefore, longitudinal studies are needed in order to understand how causal connectivity might change across recovery. Exploring correlations between concussion symptoms and causal connectivity was beyond the scope of this study. While post-concussive symptoms have been positively correlated with functional connectivity (7,17,77), correlations with causal connectivity remain underexplored and should be considered in future research. We assessed causal connectivity as a static measure, assuming it remains constant over time. However, this assumption may not accurately reflect the true nature of connectivity. Future studies should consider the dynamic nature of connectivity over time. Lastly, average values can be skewed by outliers. Future investigations should consider using median values of |*τ_i→j_*| in the group matrices.

## Conclusion

In this study, we used Information Flow Rate to estimate causal connectivity from resting-state source-reconstructed EEG in a sample of subacute concussed and healthy female athletes. To the best of our knowledge, this study is the first to evaluate causal connectivity following concussion in a female youth population. Our findings provide information regarding changes in brain network dynamics following concussion in female youth athletes and hint at a possible sex- and age-related differences in response to concussion. Understanding these changes in brain dynamics could inform personalized diagnostic and rehabilitation strategies, ultimately improving concussion management.

## Acknowledgements

We would like to acknowledge all of the athletes who participated in this study.

## Declaration of interest statement

The authors report there are no competing interests to declare.

## Funding details

This work was supported by Health Innovation Funding Investment (HIFI) Award, University of British Columbia

## Biographical note

J. McLeod (BSc) is a MSc candidate in the Department of Rehabilitation Sciences at the University of British Columbia in Vancouver, British Columbia.

S. Sattari (BEng, MASc) is a PhD student in the Department of Biomedical Engineering at the University of British Columbia in Vancouver, British Columbia.

D.T. Hristopulos (BEng, PhD) is a Professor in the School of Electrical and Computer Engineering at the Technical University of Crete in Greece.

K. Thanjavur (BEng, MEng, MASc, MSc, PhD) is a University Instructor in the Department of Physics & Astronomy at the University of Victoria in Victoria, British Columbia.

N. Virji Babul (BHSc, PT, PhD) is an Associate Professor in the Department of Rehabilitation Sciences and the Department of Physical Therapy at the University of British Columbia in Vancouver, British Columbia.

## Data availability

Datasets used in this study are available upon request to the corresponding author.

## Appendix A

All 90 connections ranked in the order of magnitude of group mean |*τ*_i→j_|.

| Control ($n=15$) | | | Concussed ($n=11$) | | |
| --- | --- | --- | --- | --- | --- |
| Transmitter | Receiver | $\vert \tau_{i \rightarrow j}\vert $ | Transmitter | Receiver | $\vert \tau_{i \rightarrow j}\vert $ |
| RC | LC | 6.54e-02 | LO | LC | 7.63e-02 |
| LT | LC | 4.83e-02 | RO | LP | 7.25e-02 |
| LP | LC | 4.61e-02 | LT | LP | 7.24e-02 |
| RP | LC | 4.58e-02 | LP | LO | 7.12e-02 |
| RP | LP | 4.58e-02 | LT | LO | 7.07e-02 |
| LC | LT | 4.44e-02 | RP | LP | 6.96e-02 |
| RT | LP | 4.24e-02 | LP | RO | 6.82e-02 |
| RO | RT | 4.10e-02 | RO | LC | 6.69e-02 |
| RO | RP | 4.05e-02 | RP | LC | 6.35e-02 |
| RT | RP | 4.01e-02 | RC | LC | 6.10e-02 |
| RP | LO | 3.84e-02 | LT | RP | 6.02e-02 |
| LP | LO | 3.81e-02 | LP | LC | 5.70e-02 |
| LP | RP | 3.58e-02 | RC | LP | 5.70e-02 |
| RP | RO | 3.47e-02 | LO | LP | 5.60e-02 |
| LT | LP | 3.45e-02 | LT | LF | 5.43e-02 |
| LT | LO | 3.43e-02 | LT | RC | 5.30e-02 |
| RF | LF | 3.42e-02 | LC | LF | 5.27e-02 |
| RO | LO | 3.37e-02 | LP | RP | 5.09e-02 |
| RC | RT | 3.34e-02 | LO | RP | 4.85e-02 |
| LC | LP | 3.28e-02 | RO | RP | 4.81e-02 |
| LP | LT | 3.27e-02 | RC | RF | 4.75e-02 |
| RT | RO | 3.25e-02 | RT | LP | 4.67e-02 |
| LP | RT | 3.19e-02 | RT | RF | 4.65e-02 |
| LO | RO | 3.15e-02 | LC | LO | 4.61e-02 |
| LO | LP | 3.12e-02 | RO | LT | 4.47e-02 |
| RP | RT | 3.08e-02 | RP | RO | 4.30e-02 |
| RT | LC | 2.97e-02 | RT | LO | 4.27e-02 |
| LF | LT | 2.95e-02 | RC | LO | 4.25e-02 |
| LC | RC | 2.88e-02 | LO | RF | 4.22e-02 |
| RC | LP | 2.75e-02 | LP | LF | 4.21e-02 |
| RC | RO | 2.65e-02 | RP | LO | 4.19e-02 |
| RP | LT | 2.64e-02 | RC | RO | 4.16e-02 |
| RF | RT | 2.63e-02 | LF | LT | 4.06e-02 |
| LC | LF | 2.62e-02 | LP | LT | 3.99e-02 |
| LF | RF | 2.60e-02 | RP | RF | 3.98e-02 |
| LT | LF | 2.59e-02 | LO | RO | 3.92e-02 |
| LO | RT | 2.55e-02 | RO | LO | 3.79e-02 |
| LP | RO | 2.53e-02 | LT | LC | 3.78e-02 |
| RP | RC | 2.49e-02 | RO | RF | 3.71e-02 |
| LO | LF | 2.44e-02 | LC | LP | 3.69e-02 |
| RF | RO | 2.43e-02 | LT | RO | 3.63e-02 |
| RT | LO | 2.43e-02 | LC | RF | 3.55e-02 |
| RC | RF | 2.40e-02 | RF | LO | 3.49e-02 |
| LO | RP | 2.39e-02 | RP | LT | 3.48e-02 |
| LC | RP | 2.37e-02 | RC | LF | 3.30e-02 |
| RC | LT | 2.32e-02 | LP | RT | 3.19e-02 |
| LP | RC | 2.27e-02 | LP | RF | 3.19e-02 |
| RF | RC | 2.26e-02 | LC | RP | 3.14e-02 |
| LO | LC | 2.23e-02 | LC | LT | 3.13e-02 |
| LF | RT | 2.22e-02 | RC | RP | 3.05e-02 |
| LC | RO | 2.19e-02 | LC | RO | 3.02e-02 |
| LO | LT | 2.18e-02 | LC | RC | 3.01e-02 |
| RF | LT | 2.15e-02 | RP | LF | 2.98e-02 |
| RT | RC | 2.14e-02 | LF | RF | 2.95e-02 |
| LC | RT | 2.12e-02 | LP | RC | 2.79e-02 |
| LT | RO | 2.10e-02 | RF | RT | 2.77e-02 |
| RO | RC | 2.01e-02 | LF | LP | 2.76e-02 |
| LC | LO | 1.96e-02 | LO | RT | 2.67e-02 |
| RC | LF | 1.93e-02 | LO | LT | 2.66e-02 |
| RO | LT | 1.89e-02 | RT | LC | 2.66e-02 |
| RT | LF | 1.88e-02 | LF | LC | 2.55e-02 |
| RT | LT | 1.82e-02 | RC | RT | 2.55e-02 |
| LT | RF | 1.80e-02 | LF | LO | 2.53e-02 |
| LF | LO | 1.75e-02 | RC | LT | 2.51e-02 |
| RP | LF | 1.69e-02 | RP | RT | 2.49e-02 |
| RO | LC | 1.69e-02 | LC | RT | 2.46e-02 |
| LT | RC | 1.69e-02 | RT | RO | 2.44e-02 |
| LC | RF | 1.65e-02 | RF | RC | 2.43e-02 |
| LO | RF | 1.64e-02 | RO | RT | 2.40e-02 |
| LT | RT | 1.64e-02 | LO | LF | 2.34e-02 |
| RO | LP | 1.61e-02 | RF | RO | 2.29e-02 |
| RC | RP | 1.56e-02 | LF | RT | 2.24e-02 |
| RC | LO | 1.55e-02 | RO | LF | 2.16e-02 |
| RT | RF | 1.53e-02 | RT | LF | 2.12e-02 |
| RP | RF | 1.50e-02 | RF | LF | 2.02e-02 |
| RO | RF | 1.50e-02 | RF | LC | 1.97e-02 |
| RO | LF | 1.50e-02 | LF | RP | 1.94e-02 |
| LF | LC | 1.48e-02 | LT | RT | 1.87e-02 |
| LO | RC | 1.45e-02 | RT | LT | 1.81e-02 |
| LF | RO | 1.38e-02 | RO | RC | 1.80e-02 |
| LP | LF | 1.37e-02 | RT | RC | 1.79e-02 |
| LT | RP | 1.37e-02 | RP | RC | 1.65e-02 |
| RF | RP | 1.34e-02 | RF | LP | 1.58e-02 |
| LP | RF | 1.33e-02 | LT | RF | 1.49e-02 |
| RF | LC | 1.30e-02 | RF | RP | 1.49e-02 |
| RF | LO | 1.28e-02 | LF | RO | 1.40e-02 |
| LF | RC | 1.24e-02 | RT | RP | 1.39e-02 |
| LF | RP | 9.69e-03 | LO | RC | 1.36e-02 |
| LF | LP | 9.67e-03 | RF | LT | 1.34e-02 |
| RF | LP | 8.89e-03 | LF | RC | 9.12e-03 |

## Notes

### Competing Interest Statement

The authors have declared no competing interest.

### Author Declarations

This study was approved by the University of British Columbia Clinical Research Ethics Board (H17-02973) in accordance with the Helsinki declarations. All participants provided written informed consent before participating.

